# The Societal Cost of Alzheimer’s Disease depends on the Diagnosis Timing

**DOI:** 10.64898/2026.01.07.26343588

**Authors:** Geoffroy Gagliardi, Nicolas Bonnet, Bertrand Schoentgen, Bénédicte Defontaines

## Abstract

**Background:** Alzheimer’s disease (AD) is a major public health issue with rising prevalence due to global aging. In France, official estimates report a societal cost of €2.3 billion, however likely underestimating the true economic burden.

**Methods:** A comprehensive economic model was developed to estimate the AD’s societal cost in France. The model incorporated diagnosis timings (early, average, late), disease stages (MCI, mild, severe AD), direct and indirect costs, and different payors.

**Results:** Total annual costs ranged from €32.5B (late diagnosis) to €36.5B (early). Early diagnosis increased total costs but reduced indirect burdens on patients. Key cost included medical expenses and home-based medical-social support. Per-patient costs varied between €62,000 (early MCI) and €175,000 (late sAD).

**Conclusion:** Earlier diagnosis increases total expenditures but improves care and cost allocation. Current and future medications focusing on the early stages of the disease, we need to invest in early diagnosis, which will reduce future expenditure.

## 1. Background

Alzheimer’s Disease (AD) and related dementia (ADRD) represent a growing challenge for healthcare systems and economies worldwide. AD, as the most common form^1,2^, is associated with substantial societal costs due to progressive cognitive decline, long disease duration, and increasing reliance on both formal healthcare services and informal caregivers. Caused by the abnormal accumulation of two proteins in the brain, i.e., amyloid and tau^3^, AD have been related to different risk factors^4^, including age with AD prevalence increasing with age. As worldwide population is growing older, the projected number of AD cases is expected to increase^2^. Consequently, economical models forecast an increasing global as well as per-patient costs over time^1^. Despite several attempts to estimate the global economic burden of AD, cross-national comparisons are limited by wide disparities in healthcare systems, long-term care funding schemes, and the availability of early diagnosis and support services. These differences highlight the need for detailed, country-level cost models, which can inform both national policies and international comparisons.

Some authors already attempted to provide better estimations, both societal and detailed, of the AD-related expenses, in different countries, overall and globally. However, the differences between healthcare systems involve adjustments and specificities making it difficult to compute an accurate model. Some models thus computed a societal cost of the disease of $958B/Y and €232B/Y for Europe alone, and projected expansion of these costs from 9.5 to 2.7-fold respectively mainly due to population ageing^5,6^.

In this paper, we present a detailed economic model aimed at providing a more accurate estimation of the societal costs of AD in France, accounting for underdiagnosis, indirect costs, and differences in care trajectories. While grounded in the French healthcare and social protection systems, our approach illustrates a methodology that could be adapted to other national contexts.

The last official epidemiological count reported 710,450 French patients diagnosed with ADRD in 2022^7^. Considering the costs assumed by the French state through the Social Security System (S3), France estimated a cost of €2.3 billion per-year (B/Y). In 2022, France official budget was €445B/Y^8^, and the total spent by the state through S3 in health was €187.6B/Y^9^. The cost of ADRD would then represent 0.53% and 1.25% of these budgets respectively. However, this information can be questioned.

First the number of patients, merging all types of dementia, only considers the diagnosed and declared patients. However, Ramaroson and colleagues estimated that only 50% of the patients suffering from AD would be diagnosed^10^. For the United States more recently, Amjad and colleagues showed that 39.5% of AD patients were undiagnosed^11^. Considering both the underdiagnosis and the prevalence of AD, some studies attempted to provide a better estimate. The resulting predictions estimated between 1.2^10^ and up to 3.6^12^ millions of patients in France. Regarding the estimated costs of the disease, this estimation only considers 1) the costs reimbursed by the S3 — i.e., not considering other payors such as private insurances or even the patients themselves —, and 2) the costs directly related to the patients and their health, i.e., leaving out indirect consequences of the disease — e.g., increased occurrence of accidents — or even the caregivers. Former literature on similar topics thus distinguished between direct and indirect costs^1^.

Finally, even considering the patients-related direct costs, this estimation only includes expenses occurring after the diagnosis, leaving aside any pathology-related costs occurring before the diagnosis, i.e., at earlier stages.

Considering these potential limits, we aimed at building a more accurate model of the yearly expenses related to AD in France. In this study, we elaborated an accounting model based on several official and scientific numbers to determine the different cost items and their payors. We computed our model considering different type of patients, in terms of which kind of treatments and funding helps they would have access to, as well as different diagnosis timing, hypothesizing that this would influence the costs as well as the stages’ duration. We hypothesized that the official estimation of the global costs of AD is greatly underestimated, even considering a number of patients lower than reality.

## 2. Methods

### 2.1. Population

According to France’s governmental official data^7^, in 2022, 710,450 patients were diagnosed with ADRD. However, based on a combination between prevalence and demographic data, Ramaroson and colleagues estimated a total of 1,198,259 patients in 2020^10^, a difference explained by the underdiagnosis with approximately 50% of patients not detected. Recently, Gabelle and colleagues made an updated model estimating the number of patients in France around 2.5 million individuals would suffer from AD^12^. For our model, we choose to use the more conservative number of 1.2M calculated by Ramaroson and colleagues.

### 2.2. Diagnosis Timing

We considered a typical patient with symptoms onset at 70 years old, and decease from the disease 14 years later. This patient would go through three clinical stages, i.e., the mild cognitive impairment (MCI), mild AD and severe AD (mAD & sAD; Figure 1).

**Figure 1.**
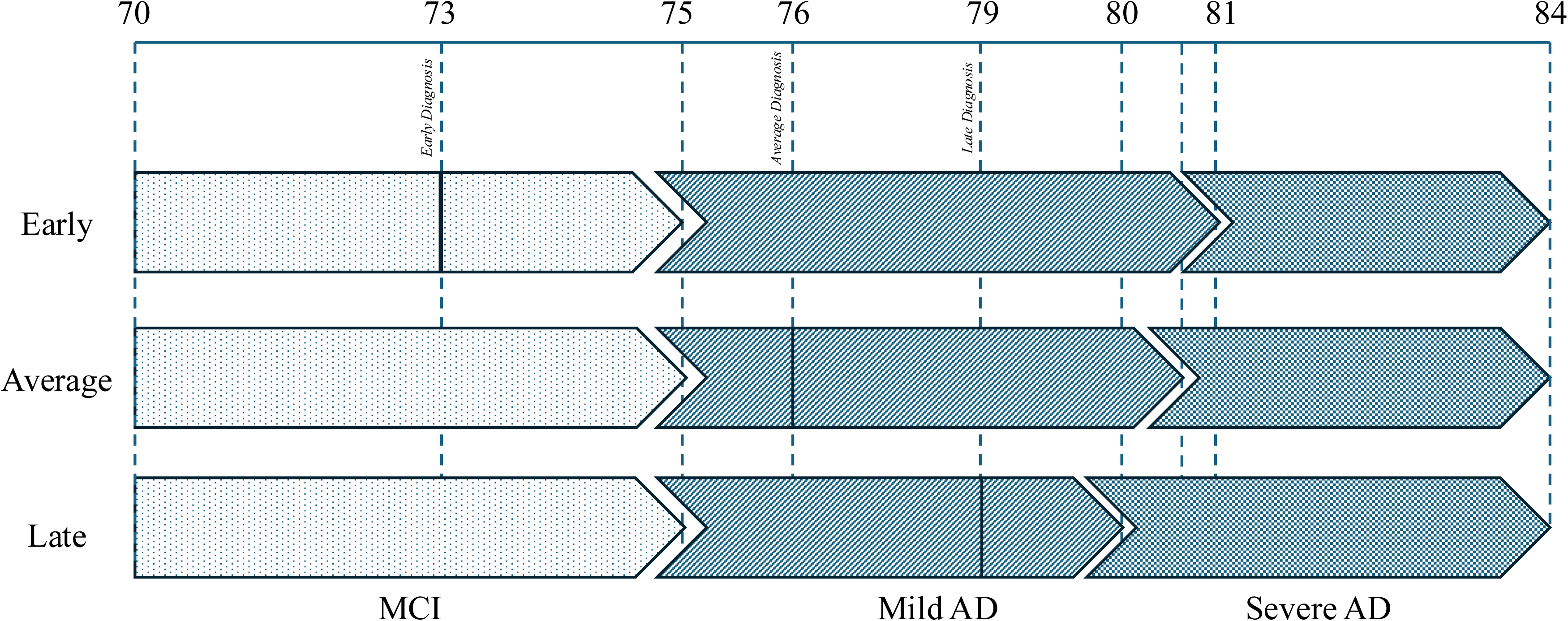
Diagnosis Timing and Stages Durations. Notes: Our model considered a typical patient with a symptom onset at 75 years old, progression from MCI to mild and then severe AD, until the patient’s decease at 84 years old. The diagnosis timing impacted the duration of the mild and sever stages, with an earlier diagnosis resulting in a longer mild AD stage.

We considered three different scenarios regarding the diagnosis timing, i.e., whether the diagnosis occurred early, late, or at a moderate timing. An early diagnosis would occur during the MCI stage, approximately 3 years after the symptoms’ onset. We considered a late diagnosis occurring approximately 4 years after progression to mAD. Finally, in our model, we considered a moderate diagnosis would occur one year after entering the mAD stage.

We considered that the stages’ duration, and particularly the progression from mAD to sAD, would vary depending on the diagnosis timing (Figure 1). The earlier the diagnosis, the longer the duration of the mAD stage, meaning that an earlier diagnosis would delay the appearance of the sAD stage. The mAD stage was thus estimated with a duration of 6, 5.5 and 5 years depending on whether the patients were diagnosed early, moderately or lately respectively.

### 2.3. Costs Items

Different cost categories have been used in literature, but there is no consensus on how to regroup these different items^1^. Literature have notably studied two main categories, i.e., Direct and Indirect costs (DC and IC). Direct costs are split into Medical (DMC) and Non-Medical costs (DNMC; Supplementary Table 1). DMC includes expenses such as health professional, hospital visits, medication or equipment^13,14^. DNMC refers to indirect medical expenses such as transportation to the clinic or home help^14^. Finally, IC are defined as invested resources without money exchange, including informal care and productivity losses^13,14^.

We also determined five different categories, i.e., Medical Expenses, Medical-Social costs occurring at Home or in Institution, the costs related to Caregivers, and finally costs related to Accidents.

The details of the considered costs, as well as the computation formula are explained in the Supplementary Methods.

### 2.4. Payor

Once the global cost of each item had been estimated for the patient at each stage, we determined the split of the expenses across different payors. Up to four actors were involved, i.e., Families, the S3, Private companies — e.g., private insurances and additional healthcare cover —, and the departmental councils.

### 2.5. Patient Profiles

Considering patients, as well as their caregivers, have different access to financial supports provided by the states and some companies, or are in different states of health, we designed nine different type profiles, split into twelve different situations (Supplementary Table 2).

The profiles were determined by a combination of both patients’ and caregivers’ states. For the patients, we considered three possible outcomes, i.e., whether the patient was receiving extensive, moderate or low treatment. We also accounted for caregivers, depending on whether they were healthy, unhealthy, or absent (i.e., no caregiver). The combination of these possibilities for patients and caregivers resulted in the nine profiles considered in our final model. Each profile was associated with a frequency, based on literature.

Depending on whether patients had access or no to certain state financial supports as well as whether they were institutionalized, we considered twelve different situations, associated with determined frequencies.

Within each of the nine profiles, we computed the costs items for each of the twelve situations. We then computed a global estimated costs for each profile and then, weighting for their frequency, we computed a global cost.

## 3. Results

### 3.1. Global Costs

We first notice that, all costs considered, less money is spent for patient when diagnosed at later stages of the disease, with a total estimated at €32.49B associated with a late diagnosis, versus €35.51B and €36.5B for an Average or Early diagnosis respectively (Table 1). That is even when considering costs items such as the occurrence of accidents. Interestingly, not only the amounts, but also the distribution of the costs vary depending on the diagnosis timing. Proportionally, the later the diagnosis, the more the expenses will rely on the patients and their families (34% for a late vs 36% for an early diagnosis), and less so to the S3 (50% vs 48% respectively; Figure 2).

**Figure 2.**
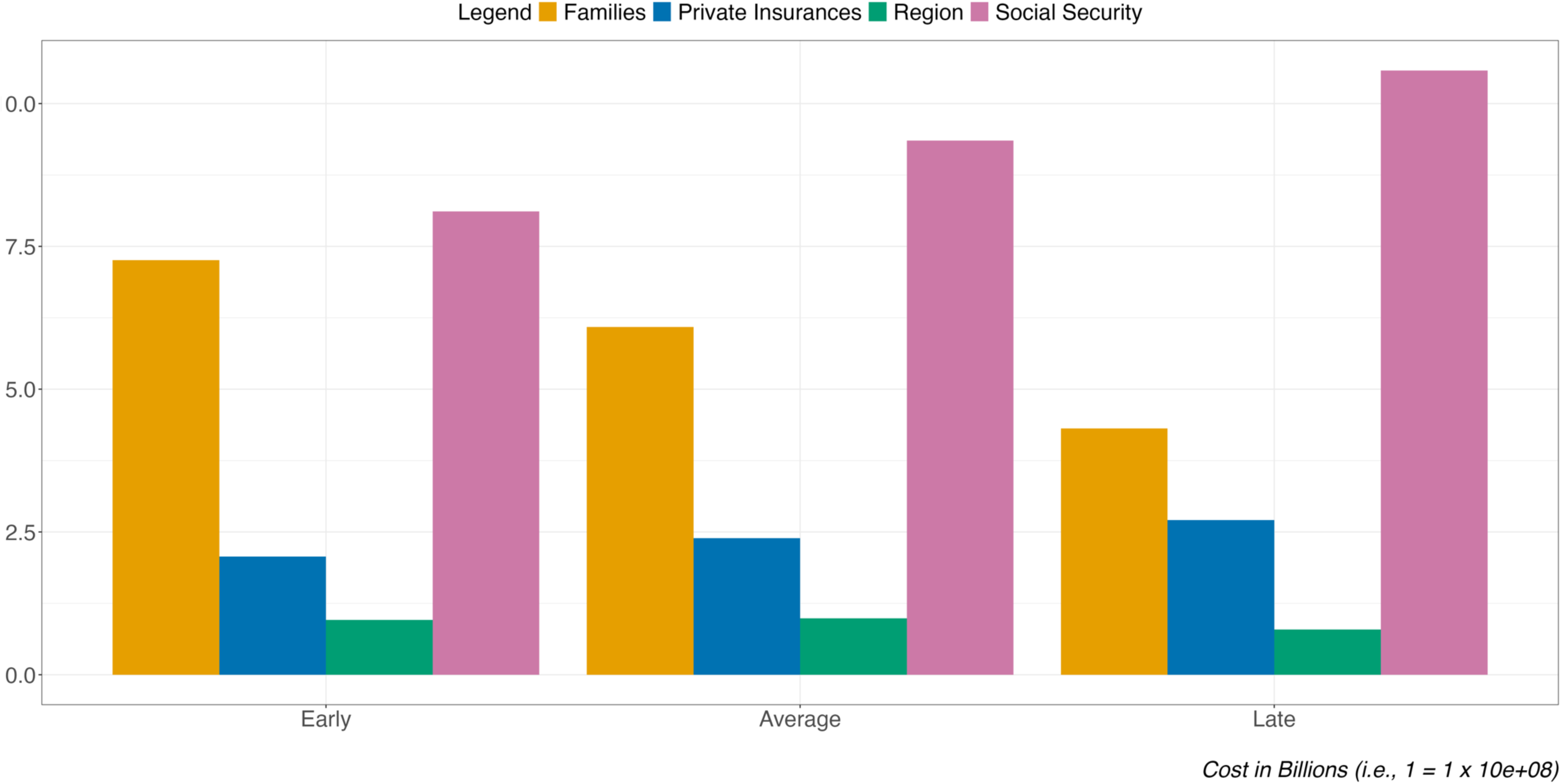
Costs repartition by payors split on the Diagnosis Timing. Notes: Costs are expressed in Billions of Euros (B€).

**Table 1.** Costs of AD split in Cost types and Time of Diagnosis. Notes: The different costs are split according to the funding source, i.e., the State through the Social Security system and the Regions, Families including the patient and their relatives, and the Private Companies through private insurances. Costs are expressed in Billions of Euros (B€). For each payor, we specified the amount as well as the percentage of the total. The total cost is split into the different types of costs, i.e., whether the expenses are Direct, medical or non-medical, or Indirect.

| Diagnosis | Cost Category | Global Cost | Families | Social Security | Private Insurances | Departmental Councils |
| --- | --- | --- | --- | --- | --- | --- |
| Early | Direct; Medical | 10,42 (29%) | 0,24 (2%) | 8,11 (46%) | 2,08 (58%) | 0 (0%) |
|  | Direct; Non-Medical | 21,51 (59%) | 9,17 (69%) | 9,05 (51%) | 1,2 (34%) | 2,09 (100%) |
|  | Indirect | 4,6 (13%) | 3,87 (29%) | 0,45 (3%) | 0,27 (8%) | 0 (0%) |
|  | **Total** | **36,53 (100%)** | **13,28 (100%)** | **17,61 (100%)** | **3,55 (100%)** | **2,09 (100%)** |
| Average | Direct; Medical | 11,97 (34%) | 0,22 (2%) | 9,35 (54%) | 2,39 (66%) | 0 (0%) |
|  | Direct; Non-Medical | 18,38 (52%) | 8,08 (64%) | 7,43 (43%) | 0,89 (25%) | 1,98 (100%) |
|  | Indirect | 5,18 (15%) | 4,37 (35%) | 0,48 (3%) | 0,32 (9%) | 0 (0%) |
|  | **Total** | **35,53 (100%)** | **12,67 (100%)** | **17,26 (100%)** | **3,61 (100%)** | **1,98 (100%)** |
| Late | Direct; Medical | 13,5 (42%) | 0,21 (2%) | 10,58 (65%) | 2,71 (73%) | 0 (0%) |
|  | Direct; Non-Medical | 12,71 (39%) | 5,64 (51%) | 5,01 (31%) | 0,54 (15%) | 1,52 (100%) |
|  | Indirect | 6,29 (19%) | 5,24 (47%) | 0,58 (4%) | 0,48 (13%) | 0 (0%) |
|  | **Total** | **32,5 (100%)** | **11,08 (100%)** | **16,17 (100%)** | **3,73 (100%)** | **1,52 (100%)** |

Overall, whether the diagnosis is obtained sooner or later, most of the costs are assumed by the S3 (from 48% to 50% depending on the diagnosis timing). The Departmental Councils was the payor with the less at charge (between 5% and 6%), followed by the Private Insurances and Companies (from 10% to 11%). No matter the timing, Families always have more than a third of the costs at their charge (from 34% to 36%).

### 3.2. Costs per Categories

Considering the model performed assuming a diagnosis occurring at the beginning of the mAD stage — labelled as “Average”—, it appeared that the most important category of cost was the DNMC (€18.38B/Y/52%; Table 2). In details, the costliest cost items were the Medical-Social costs at Home (€11.9B/33.7%) and the Medical Expenses (€12.8B/35.96%) (Supplementary Table 3). Focusing on the S3, more than half of these expenses (€9.35B/54%) were allocated to the Medical Expenses. The next most important cost category was the Medical Social at Home (€5.079B/29.43%). Regarding the patients and their families’ expenses, almost half of these were allocated to Medical Social at Home costs (€6.09B/48%), with the next most important costs being the ones dedicated to the caregivers (€3.85B/30%).

**Table 2.** Costs of AD split in Cost Categories, Time of Diagnosis and Stage of the disease. Notes: Similar from Table 2, this table adds the split of the total costs based on literature, distinguishing the Direct-Medical Costs (i.e., physician, emergency department, hospitalization, pharmacological expenses), Direct Non-Medical Costs (i.e., community care services, consumables, home modifications, institutionalization), and the Indirect Costs (i.e., lost productivity for patients and caregivers, informal care costs).

| Stage | Cost Category | Early |  |  |  |  | Average |  |  |  |  | Late |  |  |  |  |
| --- | --- | --- | --- | --- | --- | --- | --- | --- | --- | --- | --- | --- | --- | --- | --- | --- |
|  |  | Global Cost | Families | Social Security | Private Insurances | Departmental Councils | Global Cost | Families | Social Security | Private Insurances | Departmental Councils | Global Cost | Families | Social Security | Private Insurances | Departmental Councils |
| MCI | Direct; Medical | 3,688 | 0,071 | 2,888 | 0,729 | 0 | 4,411 | 0,06 | 3,481 | 0,87 | 0 | 4,411 | 0,06 | 3,481 | 0,87 | 0 |
|  | Direct; Non-Medical | 1,659 | 0,414 | 0,924 | 0,206 | 0,115 | 0,12 | 0,03 | 0 | 0,03 | 0,06 | 0,12 | 0,03 | 0 | 0,03 | 0,06 |
|  | Indirect | 1,507 | 1,2 | 0,18 | 0,127 | 0 | 1,769 | 1,462 | 0,18 | 0,127 | 0 | 1,769 | 1,462 | 0,18 | 0,127 | 0 |
|  | *Total* | *6,854* | *1,685* | *3,992* | *1,062* | *0,115* | *6,3* | *1,552* | *3,661* | *1,027* | *0,06* | *6,3* | *1,552* | *3,661* | *1,027* | *0,06* |
| Mild AD | Direct; Medical | 3,402 | 0,107 | 2,599 | 0,696 | 0 | 3,67 | 0,092 | 2,812 | 0,766 | 0 | 4,642 | 0,068 | 3,601 | 0,973 | 0 |
|  | Direct; Non-Medical | 12,336 | 5,192 | 5,361 | 0,701 | 1,082 | 9,501 | 3,894 | 4,203 | 0,518 | 0,886 | 2,605 | 0,865 | 1,328 | 0,125 | 0,287 |
|  | Indirect | 1,975 | 1,697 | 0,181 | 0,097 | 0 | 2,113 | 1,775 | 0,198 | 0,14 | 0 | 3,037 | 2,475 | 0,279 | 0,283 | 0 |
|  | *Total* | *17,713* | *6,996* | *8,141* | *1,494* | *1,082* | *15,284* | *5,761* | *7,213* | *1,424* | *0,886* | *10,284* | *3,408* | *5,208* | *1,381* | *0,287* |
| Severe AD | Direct; Medical | 3,334 | 0,062 | 2,622 | 0,65 | 0 | 3,888 | 0,072 | 3,058 | 0,758 | 0 | 4,444 | 0,083 | 3,495 | 0,866 | 0 |
|  | Direct; Non-Medical | 7,519 | 3,563 | 2,763 | 0,297 | 0,896 | 8,754 | 4,152 | 3,224 | 0,342 | 1,036 | 9,987 | 4,741 | 3,685 | 0,386 | 1,175 |
|  | Indirect | 1,113 | 0,974 | 0,09 | 0,049 | 0 | 1,299 | 1,137 | 0,105 | 0,057 | 0 | 1,484 | 1,299 | 0,12 | 0,065 | 0 |
|  | *Total* | *11,966* | *4,599* | *5,475* | *0,996* | *0,896* | *13,941* | *5,361* | *6,387* | *1,157* | *1,036* | *15,915* | *6,123* | *7,3* | *1,317* | *1,175* |
| ***Total*** |  | ***36,533*** | ***13,28*** | ***17,608*** | ***3,552*** | ***2,093*** | ***35,525*** | ***12,674*** | ***17,261*** | ***3,608*** | ***1,982*** | ***32,499*** | ***11,083*** | ***16,169*** | ***3,725*** | ***1,522*** |

The distribution of these expenses was not similar depending on the diagnosis timing. However, no matter the diagnosis timing, the most important expenses are always Medical Expenses (€10.42B/29% for early diagnosis; €13.5B/42% for late diagnosis) and Medical Social at Home (€16.1B/44%; €8.2B/25% respectively). Similarly, the greater cost categories for families are in both cases Medical Social at Home (€7.3B/55%; €4.03B/36%), while the Medical Expenses are the highest expenses for the Social Security system (€8.1B/46%; €10.6B/65%).

### 3.3. Per Patient Per Month

Regarding the Per Patient Per Month (PPPM) expenses, some constants can be reported (Table 3). To start with, for Average and Late diagnosis timings, the more severe the disease, the more expensive the estimated PPPM (sAD>mAD>MCI). This was mostly due to increases in the DNMC expenses, both the DMC and IC being relatively close from a stage to another. However, in case of an Early Diagnosis, the peak of expenses was found in the mAD stage (mAD>sAD>MCI). For equivalent stages, while the estimations were similar for the MCI stage, at the sAD stage, the PPPM was lower in case of an Early diagnosis, and peaked for the Late diagnosis.

**Table 3.**
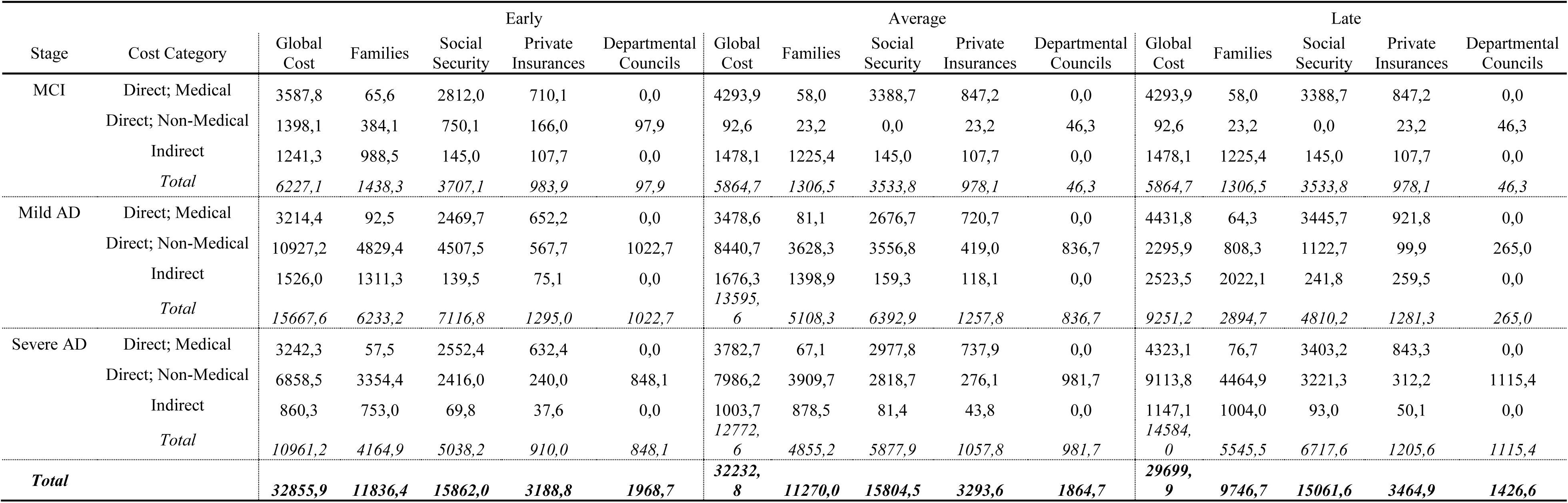
Costs of AD Per Patient Per Month (PPPM). Notes: Similar from Table 3, this table provided the Per Patient Per Month expenses split by the type of costs, diagnosis timing and disease stage.

| Stage | Cost Category | Early |  |  |  |  | Average |  |  |  |  | Late |  |  |  |  |
| --- | --- | --- | --- | --- | --- | --- | --- | --- | --- | --- | --- | --- | --- | --- | --- | --- |
|  |  | Global Cost | Families | Social Security | Private Insurances | Departmental Councils | Global Cost | Families | Social Security | Private Insurances | Departmental Councils | Global Cost | Families | Social Security | Private Insurances | Departmental Councils |
| MCI | Direct; Medical | 3587,8 | 65,6 | 2812,0 | 710,1 | 0,0 | 4293,9 | 58,0 | 3388,7 | 847,2 | 0,0 | 4293,9 | 58,0 | 3388,7 | 847,2 | 0,0 |
|  | Direct; Non-Medical | 1398,1 | 384,1 | 750,1 | 166,0 | 97,9 | 92,6 | 23,2 | 0,0 | 23,2 | 46,3 | 92,6 | 23,2 | 0,0 | 23,2 | 46,3 |
|  | Indirect | 1241,3 | 988,5 | 145,0 | 107,7 | 0,0 | 1478,1 | 1225,4 | 145,0 | 107,7 | 0,0 | 1478,1 | 1225,4 | 145,0 | 107,7 | 0,0 |
|  | Total | 6227,1 | 1438,3 | 3707,1 | 983,9 | 97,9 | 5864,7 | 1306,5 | 3533,8 | 978,1 | 46,3 | 5864,7 | 1306,5 | 3533,8 | 978,1 | 46,3 |
| Mild AD | Direct; Medical | 3214,4 | 92,5 | 2469,7 | 652,2 | 0,0 | 3478,6 | 81,1 | 2676,7 | 720,7 | 0,0 | 4431,8 | 64,3 | 3445,7 | 921,8 | 0,0 |
|  | Direct; Non-Medical | 10927,2 | 4829,4 | 4507,5 | 567,7 | 1022,7 | 8440,7 | 3628,3 | 3556,8 | 419,0 | 836,7 | 2295,9 | 808,3 | 1122,7 | 99,9 | 265,0 |
|  | Indirect | 1526,0 | 1311,3 | 139,5 | 75,1 | 0,0 | 1676,3 | 1398,9 | 159,3 | 118,1 | 0,0 | 2523,5 | 2022,1 | 241,8 | 259,5 | 0,0 |
|  | Total | 15667,6 | 6233,2 | 7116,8 | 1295,0 | 1022,7 | 13595,6 | 5108,3 | 6392,9 | 1257,8 | 836,7 | 9251,2 | 2894,7 | 4810,2 | 1281,3 | 265,0 |
| Severe AD | Direct; Medical | 3242,3 | 57,5 | 2552,4 | 632,4 | 0,0 | 3782,7 | 67,1 | 2977,8 | 737,9 | 0,0 | 4323,1 | 76,7 | 3403,2 | 843,3 | 0,0 |
|  | Direct; Non-Medical | 6858,5 | 3354,4 | 2416,0 | 240,0 | 848,1 | 7986,2 | 3909,7 | 2818,7 | 276,1 | 981,7 | 9113,8 | 4464,9 | 3221,3 | 312,2 | 1115,4 |
|  | Indirect | 860,3 | 753,0 | 69,8 | 37,6 | 0,0 | 1003,7 | 878,5 | 81,4 | 43,8 | 0,0 | 1147,1 | 1004,0 | 93,0 | 50,1 | 0,0 |
|  | Total | 10961,2 | 4164,9 | 5038,2 | 910,0 | 848,1 | 12772,6 | 4855,2 | 5877,9 | 1057,8 | 981,7 | 14584,0 | 5545,5 | 6717,6 | 1205,6 | 1115,4 |
| Total |  | 32855,9 | 11836,4 | 15862,0 | 3188,8 | 1968,7 | 32232,8 | 11270,0 | 15804,5 | 3293,6 | 1864,7 | 29699,9 | 9746,7 | 15061,6 | 3464,9 | 1426,6 |

In general, the split of the payors is found constant, with DMC being mostly taken care of by the state, while the DNMC were mainly at the charge of Families. No matter the diagnosis timing, the most important payor was always the state via the S3, followed by the families, private companies and then the regions through the department councils.

## 4. Discussion

In this study, we aimed at computing the global cost of AD in France, considering different scenarios depending on the timing of the diagnosis, the stage of the disease as well as the different levels of resource use by the patients. Comparing our results with the official number provided by the state, we found that these calculations are greatly underestimating the actual costs. This discrepancy mostly come from them only considering the direct costs reimbursed by the S3, for patients themselves — leaving aside the impacted caregivers — and only after diagnosis.

The last official estimation of governmental expenses for ADRD, in 2022, reported a €2.3B/Y for 710450 diagnosed patients^7^. However, this number is underestimating the actual amount annually spend in France for ADRD patients as both the number of patients — diagnosed or not —, the costs items and the considered payors are not representative of the actual situation. In our model, supposing an average diagnosis timing, we estimated a global cost of €35.5B/Y, with a €17.26B/Y at the charge of the state through the S3, i.e., €14.96B/Y more than the official estimation. Around the world, ADRD has been shown to be one of the costliest diseases^13^. The World Health Organization (WHO) estimated that, in 2019, 55 million patients were having dementia and, due to global population aging^15^, projected this total to raise up to 139 million by 2050^15^. Past studies have estimated a worldwide economic burden of dementia of $818B/Y in 2015, with constant progression over time through different occurrences of the model^16^. This estimation grew to $1.3 trillion level in 2019, and was anticipated to reach $2.8 trillion in 2030^15^. Focusing on European Union (EU) only, Cimler and colleagues estimated the cost of AD at €232B/Y in 2015, and projected a future total at €632.6B/Y in 2050 due to this growing number of patient and increasing per-patient costs^6^. Other authors went even further, computing the average annual costs of AD patients for different regions of EU, and estimated greater costs for Nordic regions (i.e., €43,767), followed by Southern, Western and then Eastern Europe (i.e., €35,866, €38,249 and €7938 respectively)^17^. In our study, focusing only on France, we added a supplementary layer, distinguishing the costs depending on the disease stage and the diagnosis timing (Table 3). Our results thus varied from the patient at the MCI stage, diagnosed before progressing toward dementia, costing €75725.5/Y (i.e., €6227.1/M x 12), to the patient with sAD diagnosed late associated with an annual cost of €175008.5/Y. This difference in magnitude can be explained by several factors. First, the different estimations globally include several countries and compute an average of different healthcare systems. Secondly, and for the same reason, the considered costs items may vary from a model to another. Rapp and colleagues conducted an observational studies of the AD-related costs following a cohort of patients over a 18-months period^18^. They calculated an average cost for patient ranging from €24,140 for patients diagnosed with mild AD to €44,171 for patients labeled as severe AD. These differences with our estimations can be explained in several ways. First, the sample used by the authors might represent a subsample of our considered patient types, which would necessarily impact the total estimated costs. Most importantly, this study was performed in 2018. Part of our estimated differences may then reflect the inflation (e.g., the average cost of a medical consultation raised from €26.50 to €30 in 2024) as well as some modifications in the healthcare system (e.g., reimbursement of and access to some AD-related medication for diagnosed patients)^19^.

Past studies demonstrated that with the disease progressing, the average total costs increases, with some variations from a dementia stage to another^1,20^. Our model confirms this observation, with advances stages associated with greater costs as compared to the MCI stage (Table 3). Our model again adds the detail of the diagnosis timing, showing that these average costs may vary depending on this timing, with for example, for a patient with severe dementia, the later the diagnosis, the greater the associated costs. This was due to these patients, entering later in the healthcare system, are more advanced and thus require greater resources.

These global costs can be split in subtotals. Different categories can be found in literature, and there is no absolute consensus, which makes any comparison attempt difficult^21^. Based on literature, we organized our cost reports in two main categories, i.e., DC and IC, with DC split into DMC and DNMC. Taken separately, the DMC, DNMC and IC were globally estimated by the World Health Organization at $3624, $13,128 and $12,393 per patient per year^22^. Focusing on European countries, studies have reported DMC varying from €171 to $313 per patient per month (PPPM)^1^. Our estimation, depending on the diagnosis timing and the disease stage, ranged from €43,053.2/Y for early MCI to €175,008.2/Y for severe AD with a late diagnosis. Aside from the 22 years separating our studies that could explain part of the discrepancy, other factors could also contribute. For example, the time spent by the caregiver for the patient, which was found significantly varying from a country to another^1^, which would impact the average done when considering together multiple countries.

In 2018, Rapp and colleagues estimated the total direct costs PPPM over 18 months in France and found expenses ranging from €1,341 to €2454 depending on the stage. Split in DMC and DNMC, they estimated the PPPM to range from €285 to €451 and €359 to €742 respectively. In our model, our estimations ranged from €3,587.8 to €4,323.1 and €1,398.1 to €9,113.8 respectively. As we can see, and for similar reasons as mentioned before, our estimations are more important than these of these authors, reflecting both methodological discrepancies as well as the important increase of AD-related expenses.

To note, in our model, Early diagnosis was not associated with increased PPPM costs as the disease progresses, with more expenses found at the mAD stage, followed by the sAD and then the MCI stages. Far from opposing to past literature, this is most certainly due to the calculation method. In our model, the costs were computed at each stage, and then weighted depending on the stage duration in years, and divided to obtain the PPPM. Considering that, for Early diagnosis, the sAD stage was the shortest and the mAD the longest, this could be the reflection of this calculus.

In general, and as we discussed above, the disease progression is associated with greater costs, as shown by both our model and previous calculations^1,20,23^. This is consequential to increased costs for the patients, as well as their caregivers whom burden increases as the disease progresses^20^. Global population growing older, with an estimation of 1/3 French older than 60 years in 2050^19^, the number of patients with ADRD will significantly increase^5,17^. Currently, the anti-Alzheimer’s medications tested and/or released targets patients in early stages^3^, inducing an incentive to any initiative allowing earlier detection or reducing the duration of later cases, which could translates into costs savings^1,6^. Past studies have even showed that prolongating the duration of the mAD stage of the disease would lead to more savings than prolongating the Moderate stage duration^6^. Another option, rather than delaying the progression from a stage to another, is to focus on strategies that might delay the onset of the disease itself. As an example, Zissimopoulos and colleagues estimated and increase of +153% of the number of individuals above 70 years old from 2010 to 2050^5^. In their model, this was associated with an annual global cost increase from $307 billion to $1.5 trillion. They then explained that delaying the delaying the onset of AD for 5 years would lead to lower the global prevalence and the costs of 41% and 40% respectively, with an estimated $599 billion savings. As our understanding and awareness of the disease progresses, diagnoses will be made earlier. With all the advances being made, we can expect our ability to prolongate further the early stages duration, thereby reversing this cost trend.

### 4.1. Limitations

Some limitations can be mentioned regarding our model construct and results. First, in our model, it is posted that every patient will obtain a diagnosis at some point. However, we know that it is not always and necessarily the case. Considering this, even if low, proportion of patient who will never be diagnosed could reinforce the precision of our model. Another limitation regards the patient profiles. Literature have shown that some demographic variables — e.g., age, sex, education, belonging to minorities or presence/absence of comorbidities — have a significant effect on costs^13,20,24^. This could notably be explained by patients’ resource use or access^13^. These variables have not been considered in the present version of our model. Future version could benefit from including them to better reflect their effect.

### 4.2. Conclusion

AD, and neurodegenerative diseases in general, are a global public health issue. Epidemiological and prospective data show that an ever-increasing proportion of the population is or will be affected. It should be noted that, at present, most of the costs considered in our model concern patients after diagnosis. After decades of setbacks, pharmacological research is now largely focused on early stages of the disease. Today, we feel it is important to reflect on the allocation of our resources, and to invest significantly in the examination and management of these early stages. In our view, early detection and diagnosis of the disease, along with current and future developments in research on delaying the disease onset or early stages duration, would lead to better patient care and, ultimately, optimization of resources, both economically and in terms of care.

## Data Availability

All data produced in the present study are available upon reasonable request to the authors

## Acknowledgments

We thank the consulting firm Kea & Partners, based in Paris, for their support in developing the model on which the conclusions of this article are based. We also thank the international organization Ashoka for facilitating this collaboration and enabling the pro bono contribution of Kea & Partners.

## Conflicts of Interest

Authors report no conflict of interest.

## Fundings Sources

This research received no specific grant from any funding agency in the public, commercial, or not-for-profit sectors.

## Consent Statement

As the data implemented in the model all come from official and/or scientifically published data in order to construct “typical patients”, no informed consent was required for this study. No patients and/or caregivers were interviewed as part of this research to feed the model.

## Authorship

All named authors meet the International Committee of Medical Journal Editors (ICJME) criteria for authorship for this article, take responsibility for the integrity of the work, and have given their approval for this version to be published.

## Authors Contributions

All authors have contributed to the writing of the manuscript. GG performed the initial literature review and writing of the manuscript. NB designed the first version of the economic model. GG and NB designed the last model version, curated the base values included for the direct and indirect costs, and performed the computation of the model. BS and BD contributed as consulted clinician experts for some of the used values in the model and supervised the overall project.

## Data accessibility

For this work and based both on our hypotheses and official numbers, a whole dataset has been created. This complete data set, along with the complete set of formula and calculi, is freely available on GitHub at the following link: https://github.com/GagGeo/eco-ad.

### Used Abbreviations

AD: Alzheimer’s Disease;
ADRD: AD and related diseases;
mAD/sAD: Mild/Severe AD;
DC: Direct Costs;
DMC: Direct Medical Costs;
DNMC: Direct Non-Medical Costs;
IC: Indirect Costs;
B/T: Billion/Trillion.

## SUPPLEMENTARY TABLES

**Supplementary Table 1.**
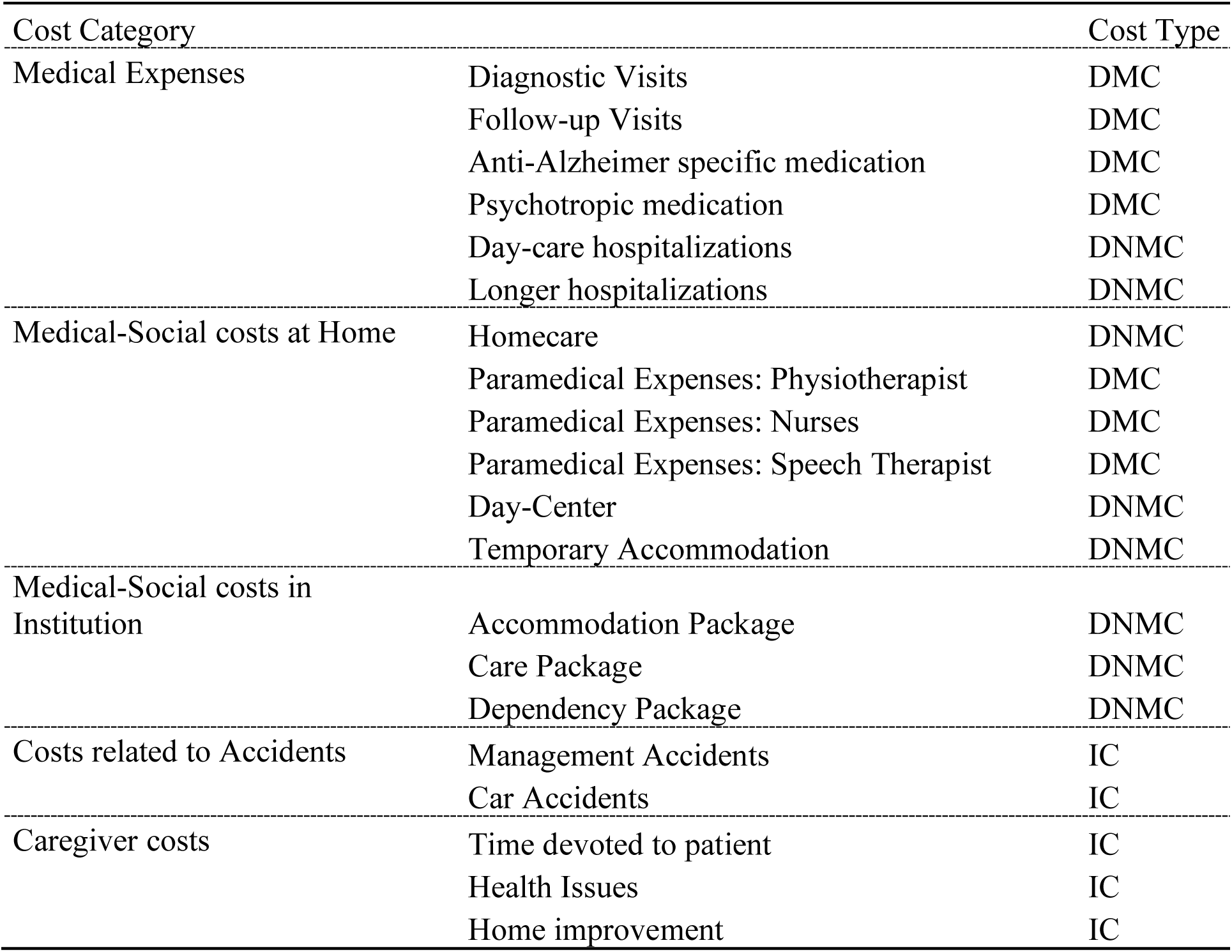
Cost Types Categories. Notes: DMC = Direct Medical Costs; DNMC = Direct Non-Medical Costs; IC = Indirect Costs

**Supplementary Table 2.**
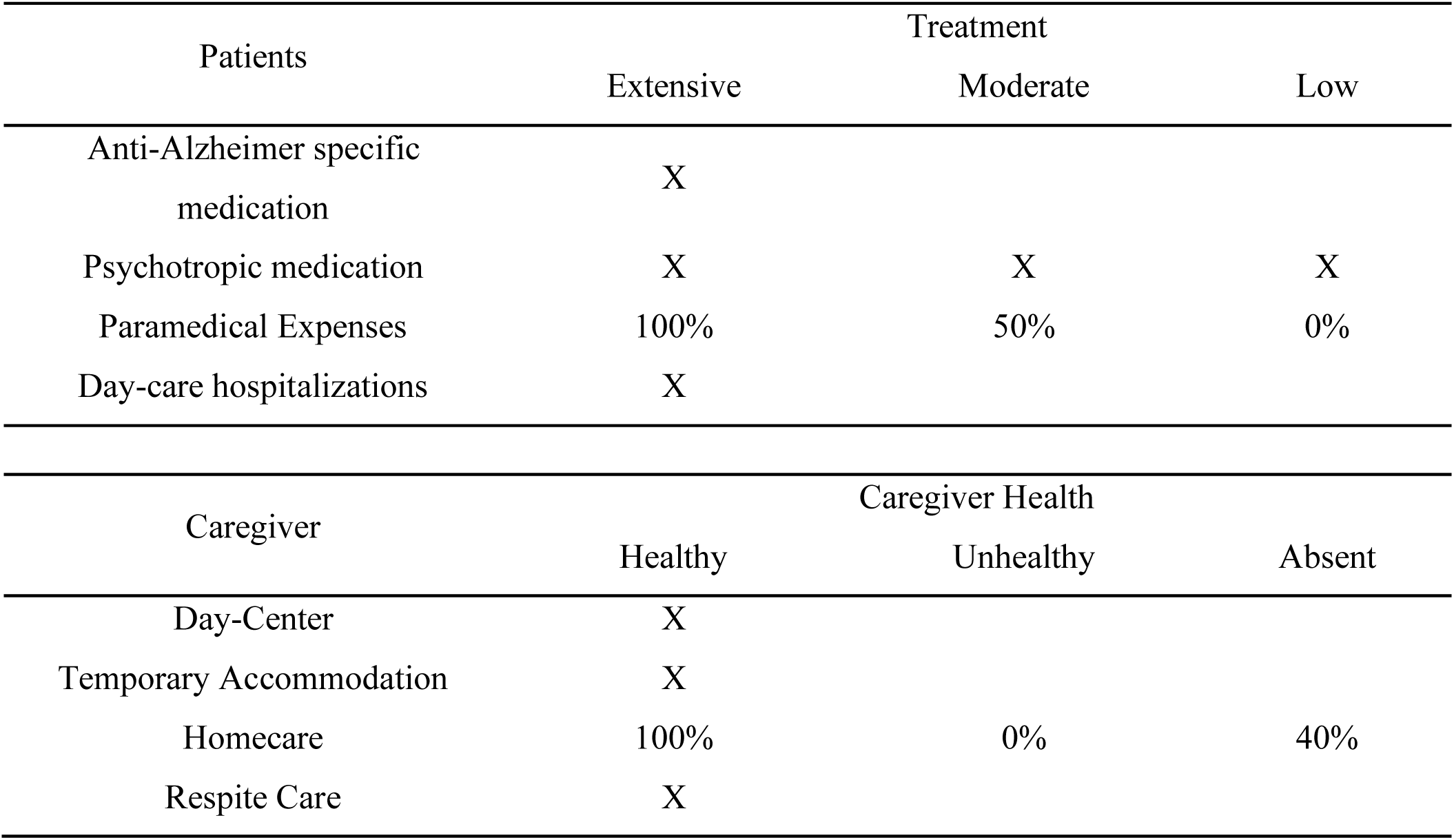
Patients and Caregiver Profiles, Resources Access.

**Supplementary Table 3.**
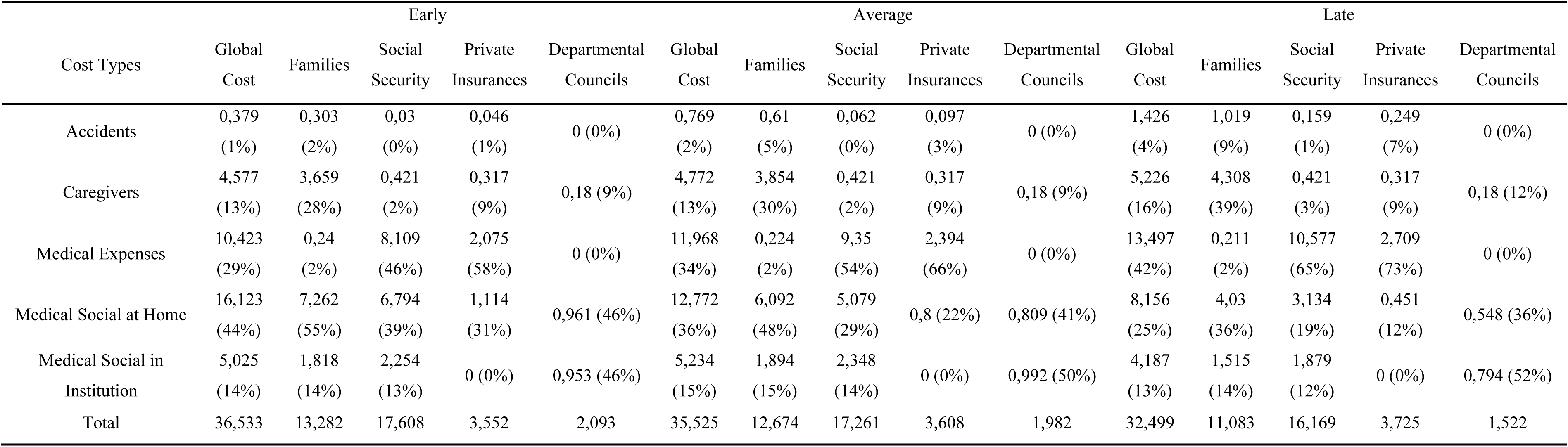
Costs of AD split in Cost types and Time of Diagnosis. Notes: Similar from Table 1, this table adds the split of the total costs into the different types of costs, i.e., whether the expense occurs from medical costs, medical and social costs at home or in institution, the increased risks of accidents or related to caregivers.

## SUPPLEMENTARY METHODS: Costs Items Details

### Medical Expenses

Medical expenses were distributed between consultations, medication and hospital costs.

For the consultations, we included both the diagnostic and the follow-up consultations, each having different costs depending on the specific exams required, especially for the diagnosis. While the follow-up consultations were only counted considering the cost of a single visit at a primary care provider or a specialist — i.e., neurologist or psychiatrist —, the diagnostic consultations included in addition confirmatory exams such as the neuropsychological examination, brain imaging or lumbar punction. The single cost of a follow-up consultation was then multiplied by the estimated number of occurrences per-year depending on the stages and for the duration of the stages.

For the medication, we included the Anti-Alzheimer specific medication, and the Psychotropic medication. Currently, eight different Anti-Alzheimer drugs (AAD) can be listed, each acting on different mechanisms, used at different stages and for different purposes^2^. In our model, we focused on the drugs aiming at treating the cognitive symptoms of AD and recommended for the clinicians in France. These were the Donepezil, the Rivastigmine and the Memantine. By acting on the neurotransmitter levels, these medications only have a symptomatic effect, and they do not intend to slow down or stop the disease’s evolution or pathological underlying cause. We average the different costs of these medications to obtain a per-day cost and multiplied it by the days-per-year average prescription for the duration of the stages.

Other than AD-related cognitive symptoms and decline, extensive literature have shown the presence of neuro-psychiatric symptoms in AD^25^. These may include symptoms such as agitation, depression, delusions and others^26^. Among the available treatments, the Risperdone is one of the most used, having shown convincing proofs of efficacy^27^. For our model, we considered the average daily cost of the medication for a standard duration of 42 days per year maximum. This was then computed for the duration of the stages, starting at the mAD stage.

Finally, hospitalization costs were split between day-care and longer hospitalizations. The cost of hospitalizations was obtained by multiplying the daily rate by the average duration of hospitalization depending on age, adding a supplementary risk factor associated with the presence of AD. For day-care, we considered the fixed cost of a day-care once a week.

### Medical-Social costs at Home

For the home-related medical-social costs, we included different types of expenses among which the homecare and paramedical expenses, as well as day center and temporary accommodation costs.

Homecare corresponds to employment of housekeepers, home helps, meal deliveries, etc. We computed the estimated time of helps and number of meals delivered, considering their unitary cost, for the duration of the stages.

Paramedical expenses cover the use of professionals such as physiotherapists, nurses or speech therapists, coming at home to help or stimulate the patient.

Finaly, we considered the costs of day-center and temporary accommodation, such as provided in some medicalized retirement houses. The costs and needs were both fixed numbers, and the variations we computed on the duration of the stages.

### Medical-Social costs in Institution

Institutionalized patients are presented with a global bill including three different services, split in the care, dependency and the accommodation packages. For the current model, we considered each package independently as each have different payor repartitions.

### Caregiver costs

Along with the patients’ disease-related cost, their decline as well as its consequences have been shown to have direct consequences on the caregivers. Caregivers can exhibit health issues such as psychiatric syndromes, cardiovascular symptoms and other health issues^13^. In our model, we computed the costs related to the caregiver’s health condition, as well as the time devoted to the patient — included as a loss of income and then an indirect societal cost. We also included the costs related to necessary home improvement to adapt to the patients’ decline.

### Costs related to Accidents

The presence of a neurodegenerative disease may lead to different kind of errors and accidents. We considered both car and management accidents for the present model.

Management accidents referred to all the mishaps — e.g., poor financial decisions, wasteful spending, scams — that can occur and be increased by the presence of the cognitive symptoms associated with AD. We estimated an average cost — i.e., 1000€ — and considered an average number of such accidents at approximately one per year.

Regarding car accidents, we first computed the average cost of accidents using the official estimates for material and personal costs associated to accidents ^A^. These costs were associated with a prevalence, both raw number of accidents as well as the proportion of these accidents involving personal injuries. We then added the increased risk of accidents associated with AD. Following previous published estimations, we considered an increased risk of car accidents of x2 for the MCI stage ^B^, and of x7.9 for the mild-AD without diagnosis ^C^. The mAD post diagnosis and sAD were not considered at risk as we posed that they would not engage into driving. This was computed for the duration of the stages.

### Costs Items Categories

Costs items were split into the DMC / DNMC / IC categories. DMC included the medical expenses (consultations, medication and hospital costs). DNMC included the Medical Social costs, both at home and in institution. Finally, IC included the costs related to Accidents and the caregivers.

## Notes

### Competing Interest Statement

The authors have declared no competing interest.

### Funding Statement

This study did not receive any funding

