## Supplemental Tables for "The Societal Cost of Alzheimer’s Disease depends on the Diagnosis Timing"

### *SUPPLEMENTARY TABLES*

*Supplementary Table 1.* Cost Types Categories
Notes: DMC = Direct Medical Costs; DNMC = Direct Non-Medical Costs; IC = Indirect Costs

*Supplementary Table 2*. Patients and Caregiver Profiles, Resources Access

*Supplementary Table 3. Costs of AD split in Cost types and Time of Diagnosis.*
Notes: Similar from Table 1, this table adds the split of the total costs into the different types of costs, i.e., whether the expense occurs from medical costs, medical and social costs at home or in institution, the increased risks of accidents or related to caregivers.

Supplementary Table 1. Cost Types Categories

| Cost Category | | Cost Type |
|---|---|---|
| Medical Expenses | Diagnostic Visits | DMC |
| | Follow-up Visits | DMC |
| | Anti-Alzheimer specific medication | DMC |
| | Psychotropic medication | DMC |
| | Day-care hospitalizations | DNMC |
| | Longer hospitalizations | DNMC |
| Medical-Social costs at Home | Homecare | DNMC |
| | Paramedical Expenses: Physiotherapist | DMC |
| | Paramedical Expenses: Nurses | DMC |
| | Paramedical Expenses: Speech Therapist | DMC |
| | Day-Center | DNMC |
| | Temporary Accommodation | DNMC |
| Medical-Social costs in Institution | Accommodation Package | DNMC |
| | Care Package | DNMC |
| | Dependency Package | DNMC |
| Costs related to Accidents | Management Accidents | IC |
| | Car Accidents | IC |
| Caregiver costs | Time devoted to patient | IC |
| | Health Issues | IC |
| | Home improvement | IC |

Supplementary Table 2. Patients and Caregiver Profiles, Resources Access

| Patients | Treatment | | |
|---|---|---|---|
| | Extensive | Moderate | Low |
| Anti-Alzheimer specific medication | X | | |
| Psychotropic medication | X | X | X |
| Paramedical Expenses | 100% | 50% | 0% |
| Day-care hospitalizations | X | | |

| Caregiver | Caregiver Health | | |
|---|---|---|---|
| | Healthy | Unhealthy | Absent |
| Day-Center | X | | |
| Temporary Accommodation | X | | |
| Homecare | 100% | 0% | 40% |
| Respite Care | X | | |

Supplementary Table 3. Costs of AD split in Cost types and Time of Diagnosis.

| | Early | | | | | Average | | | | | Late | | | | |
|---|---|---|---|---|---|---|---|---|---|---|---|---|---|---|---|
| Cost Types | Global Cost | Families | Social Security | Private Insurances | Departmental Councils | Global Cost | Families | Social Security | Private Insurances | Departmental Councils | Global Cost | Families | Social Security | Private Insurances | Departmental Councils |
| Accidents | 0,379 (1%) | 0,303 (2%) | 0,03 (0%) | 0,046 (1%) | 0 (0%) | 0,769 (2%) | 0,61 (5%) | 0,062 (0%) | 0,097 (3%) | 0 (0%) | 1,426 (4%) | 1,019 (9%) | 0,159 (1%) | 0,249 (7%) | 0 (0%) |
| Caregivers | 4,577 (13%) | 3,659 (28%) | 0,421 (2%) | 0,317 (9%) | 0,18 (9%) | 4,772 (13%) | 3,854 (30%) | 0,421 (2%) | 0,317 (9%) | 0,18 (9%) | 5,226 (16%) | 4,308 (39%) | 0,421 (3%) | 0,317 (9%) | 0,18 (12%) |
| Medical Expenses | 10,423 (29%) | 0,24 (2%) | 8,109 (46%) | 2,075 (58%) | 0 (0%) | 11,968 (34%) | 0,224 (2%) | 9,35 (54%) | 2,394 (66%) | 0 (0%) | 13,497 (42%) | 0,211 (2%) | 10,577 (65%) | 2,709 (73%) | 0 (0%) |
| Medical Social at Home | 16,123 (44%) | 7,262 (55%) | 6,794 (39%) | 1,114 (31%) | 0,961 (46%) | 12,772 (36%) | 6,092 (48%) | 5,079 (29%) | 0,8 (22%) | 0,809 (41%) | 8,156 (25%) | 4,03 (36%) | 3,134 (19%) | 0,451 (12%) | 0,548 (36%) |
| Medical Social in Institution | 5,025 (14%) | 1,818 (14%) | 2,254 (13%) | 0 (0%) | 0,953 (46%) | 5,234 (15%) | 1,894 (15%) | 2,348 (14%) | 0 (0%) | 0,992 (50%) | 4,187 (13%) | 1,515 (14%) | 1,879 (12%) | 0 (0%) | 0,794 (52%) |
| Total | 36,533 | 13,282 | 17,608 | 3,552 | 2,093 | 35,525 | 12,674 | 17,261 | 3,608 | 1,982 | 32,499 | 11,083 | 16,169 | 3,725 | 1,522 |
