## Supplemental Methods for "The Societal Cost of Alzheimer’s Disease depends on the Diagnosis Timing"

### *Supplementary References*

A. Observatoire national interministériel de la sécurité routière. La sécurité routière en France. Bilan de l’accidentalité de l’année 2021. Paris: Ministère de l’Intérieur, 2022.

B. Retchin SM, Hillner BE. The Costs and Benefits of a Screening Program to Detect Dementia in Older Drivers. Med Decis Making 1994; 14: 315–24.

C. Friedland RP, Koss E, Kumar A, et al. Motor vehicle crashes in dementia of the alzheimer type. Ann Neurol 1988; 24: 782–6.
